# Incidence and outcomes of healthcare-associated COVID-19 infections: significance of delayed diagnosis and correlation with staff absence

**DOI:** 10.1101/2020.07.24.20148262

**Authors:** Kirstin Khonyongwa, Surabhi K. Taori, Ana Soares, Nergish Desai, Malur Sudhanva, Will Bernal, Silke Schelenz, Lisa A. Curran

## Abstract

**Background:** The sudden increase in COVID-19 admissions in hospitals during the SARS-CoV2 pandemic of 2020 has led to onward transmissions among vulnerable inpatients.

**Aims:** This study was performed to evaluate the prevalence and clinical outcomes of Healthcare-associated COVID-19 infections (HA-COVID-19) during the 2020 epidemic and study factors which may promote or correlate with its incidence and transmission in a London Teaching Hospital Trust.

**Methods:** Electronic laboratory, patient and staff self-reported sickness records were interrogated for the period 1^st^ March to 18^th^ April 2020. HA-COVID-19 was defined as symptom onset >14d of admission. Test performance of a single combined throat and nose swab (CTNS) for patient placement and the effect of delayed RNA positivity (DRP, defined as >48h delay) on patient outcomes was evaluated. The incidence of staff self-reported COVID-19 sickness absence, hospital bed occupancy, community incidence and DRP was compared HA-COVID-19. The incidence of other significant hospital-acquired bacterial infections (OHAI) was compared to previous years.

**Results:** 58 HA-COVID-19 (7.1%) cases were identified. As compared to community-acquired cases, significant differences were observed in age (p=0.018), ethnicity (p<0.001) and comorbidity burden (p<0.001) but not in 30d mortality. CTNS negative predictive value was 60.3%. DRP was associated with greater mortality (p=0.034) and 34.5% HA-COVID-19 cases could be traced to delayed diagnosis in CA-COVID-19. Incidence of HA-COVID-19 correlated positively with DRP (R=0.7108) and staff sickness absence (R=0.7815). OHAI rates were similar to previous 2 years.

**Conclusion:** Early diagnosis and isolation of COVID-19 would help reduce transmission. A single CTNS has limited value in segregating patients into positive and negative pathways.

## Introduction

The COVID-19 [SARS-CoV-2] outbreak started in China in late December 2019 and was eventually declared a Public Health Emergency of International Concern on 30^th^ January, 2020. The first cases identified in the UK were in international travellers but by February, local transmission was observed. London experienced the largest number of cases recorded in any UK region (1) while a large part of the total burden of disease was in South-East London where this study was performed.

To increase capacity for intensive care of severely ill COVID-19 patients in the Hospital Trust studied, elective work was minimised. Eventually specific wards and intensive care units (ICUs) became cohort areas for affected patents though non COVID-19 emergency work continued. Due to the high prevalence of infection during the peak of the outbreak, one of the suggested strategies to prevent healthcare transmission was to screen all patients on admission by a single combined nose and throat swab assessed for SARS-CoV-2 RNA to allow segregation into COVID-19 positive and non COVID-19 cohort wards.

Recent publications have identified advanced age, comorbidities and male gender among as major risk factors for severity and mortality in COVID-19 (2, 3) and the impact of ethnicity is being explored (4). Healthcare associated COVID-19 infections (HA-COVID-19) have been reported in other studies (5) but the literature on epidemiology, risk factors and outcomes of healthcare acquisition of COVID-19 among healthcare inpatients is lacking.

This study was performed to determine the burden, risk factors and clinical outcomes of adult HA-COVID-19 infections and evaluate factors which may promote or correlate with the incidence and transmission of HA-COVID-19. The latter included assessment of the utility of a single combined throat and nose swab (CTNS) for patient placement, delayed RNA positivity, COVID-19 patients as sources of infection, self-reported COVID-19 sickness absence among hospital staff hospital bed occupancy, community incidence, and the incidence of other significant hospital-acquired infections.

## Methods

### Setting

This study was conducted at the main hospital site of a large tertiary care teaching hospital in south-east London from 1^st^ March to 18^th^ April 2020. Before the COVID-19 outbreak, the capacity was 960 beds including 73 adult intensive care beds. There is a mix of specialities including haemato-oncology including haematopoetic stem cell transplant and CAR-T therapy, liver transplant, neurosurgery, neurology, women’s health, paediatric, renal, respiratory and endocrinology. The hospital serves a socioeconomically deprived region of London.

All patients who were SARS-CoV-2 RNA positive from a respiratory sample and who were admitted to hospital for at least an overnight stay were included in the study.

### Case definitions

#### COVID-19 infection

either clinical or radiological evidence of pneumonia or acute respiratory distress syndrome, or influenza-like illness with fever ≥37.8°C and acute onset of at least one of the following: persistent cough (with or without sputum), hoarseness, nasal discharge or congestion, shortness of breath, sore throat, wheezing, sneezing and PHE estimate of the incubation period (range 1 to 14 days, median 5 days) (6, 7).

Community-associated COVID-19 infection (CA-COVID-19):

1. All symptoms on admission in keeping with above symptoms of COVID-19 AND
2. Respiratory sample positive for SARS-CoV-2 RNA

Hospital-associated COVID-19 infection (HA-COVID-19):

1. An alternative proven aetiology for all presenting symptoms on admission (non COVID-19) AND
2. Symptoms of COVID-19 developed >14 days after admission AND
3. Respiratory sample positive for SARS-CoV-2 RNA >14 days of admission

Indeterminate acquisition:

1. Proven alternate aetiology for all presenting symptoms on admission AND
2. COVID-19 symptom developed within 48h but <=14 days after admission AND
3. Respiratory sample positive for SARS-CoV-2 RNA

Among the indeterminate cases those presenting from 8-14d after admission were designated late indeterminate.

#### Asymptomatic COVID-19

Patients who did not have any symptoms of COVID-19 14d after SARS-CoV-2 RNA positive result or up to the time of discharge.

NHS England released its reporting criteria in May 2020 (written communication described in supplementary data) following which cases were also classified as per date of the SARS-CoV-2 RNA detection. We compared our classification with this to determine any differences which may be associated with using the latter methodology (see supplementary information).

### Infection prevention and control (IPC) measures

All patients suspected of having a respiratory illness were seen at the emergency department and triaged for admission. If inpatient care was required, they were admitted to a ward designated as a holding area (18 en-suite single rooms) while awaiting results of investigations. If a swab was SARS-CoV-2 RNA positive, then they were kept in a side room with IPC precautions or in a designated COVID-19 cohort ward/ICU. If the swab was SARS-CoV-2 RNA negative, they were placed with other non-COVID-19 patients.

Initially, respiratory samples were sent to the Virus Reference Department, Public Health England Colindale for SARS-CoV-2 RNA testing. Local, SARS-CoV-2 RNA by real time PCR testing was introduced on 29th February 2020. This reduced the mean turnaround time of SARS-CoV-2 RNA results from receipt in the laboratory to less than 6 hours which enabled rapid movement of patients from within the holding area. When the requirement for beds with ventilator support increased, other areas of the hospital were repurposed to accommodate 102 ICU beds.

Negative pressure rooms were limited in the hospital and a risk assessment was done to reduce the impact of aerosols. A separate area for donning and doffing personal protective equipment was established for each ward. When side room availability reached capacity, parts of the ward were created as cohort areas with bed spaces segregated from the other, curtains always to be closed, and where possible, allotted a separate toilet and dedicated nurse. Staff members were advised to wear (personal protective equipment (PPE) and FFP3 masks as appropriate. Training and mask fit testing sessions were organised in preparation to the expected outbreak from February 2020 and continued throughout the period of the outbreak. Public Health England guidance and updates were followed for all other aspects of infection prevention and control.

Cleaning of environmental surfaces and clinical equipment was implemented as per PHE recommendations. Curtains were changed if a non COVID-19 patient was to be admitted to the bed space vacated by a COVID-19 patient.

When an HA-COVID-19 case was identified, actions included enhanced training for correct PPE usage, rapid transfer of patients to a COVID-19 positive cohort ward, deep cleaning followed by enhanced cleaning frequency till no further transmission was seen.

### Virological methods

A combined throat and nose a swab (CTNS) in VTM or respiratory fluid such as bronchoalveolar lavage (BAL) were assessed using RdRp gene assay (PHE Colindale or locally after 29 Feb 2020) for SARS CoV-2 RNA. Local interpretation of PCR curves was performed using PCR: AI machine-learning software (8)

### Clinical, laboratory and outcome data

SARS-CoV-2 RNA results data was extracted from the laboratory information management system (WinPath). Clinical and demographic details were extracted from electronic patient records (EPR and ICCA). Age, gender and ethnicity (as Black Asian and Minority Ethnicities (BAME) or non-BAME), chronic kidney disease (CKD), hypertension, malignancy, dementia, chronic obstructive pulmonary disease (COPD), diabetes and the Charlson Comorbidity Index (CCI-age adjusted) were noted. Patients without electronic clinical records were excluded. Patients were followed up for 30 days. Duration of hospital stay, readmission after discharge and 7, 14 and 30d mortality were recorded. ICU admission within 7 days of COVID-19 diagnosis was used as a marker for severe disease and a subset analysis was performed.

### Test performance of single CNTS

True positives were all CA-COVID-19 cases positive on the first CTNS (taken within 48h of admission), true negatives were patients with a negative CTNS swab within 48h of admission and had a minimum stay of 14days and SARS-CoV-2 RNA negative or diagnosed within this admission. False positives were none (assuming the PCR test does not give false positives) and false negatives were all CA-COVID-19 cases negative on the first swab (within 48h).

<u>Delayed SARS-CoV-2 RNA positivity</u> was defined as

1. No RNA positive results within 48h of presentation (for CA-COVID-19) OR
2. No RNA positive results within 48h of symptom onset (for indeterminate and HA-COVID-19).

The outcomes of patients with a delayed RNA positive result were compared with those without a delay.

### Source and incubation period of HA-COVID-19 and late indeterminate cases

If an index case patient was found in the same ward as a HA-COVID-19 case (within 14d prior) they were recorded as a potential source. Duration of exposure of each HA-COVID-19 case to a known positive patient was calculated and the incubation period was determined using the midpoint of the exposure period up to the development of symptoms and expressed as a range from the earliest and latest contact with the known positive. Asymptomatic cases were excluded from this analysis.

### Staff self-reported absence

Workforce Development database (Health-Roster) was interrogated for staff absences due to 1: COVID-19, 2: cold, cough and flu-like illness, 3: chest and respiratory problems. This did not include those self-isolating or shielding as per UK government advice (9). Absences due to 2 and 3 above from 2019 were evaluated for comparison. Staff was grouped as per patient facing roles (nurses, doctors, additional clinical services and allied health professionals), non-patient facing, high nosocomial exposure risk (estates and ancillary, Scientific and Technical and Healthcare Scientists) and non-clinical (Administrative and Clerical) to compare association with healthcare contact. Only one episode of sickness was recorded per 14d period for each staff member. Staff members deployed to other roles were excluded. Records for staff not directly employed by the Trust were not available for analysis e.g. porters and catering.

### Hospital bed occupancy data, community incidence and other significant healthcare-associated bacterial infections (HAB)

These included bloodstream infections due to *Staphylococcus aureus* (MRSA and MSSA), *E*.*coli*, Klebsiella spp, *Pseudomonas aeruginosa*, and vancomycin-resistant Enterococci and toxigenic *Clostridioides difficile* infections detected after 48h of admission. Their incidence from January to May 2020 was compared to their average incidence from 2018 and 2019 (Jan to May). Results are in figure S4 (supplementary information). Bed occupancy was derived from the Business Intelligence Unit of the Hospital and community incidence was derived from national population data (10) and Department of Health reports of COVID-19 cases (1)

### Statistical methods

Categorical variables were compared using the chi square test or Fisher’s exact test. Continuous variables not normally distributed were presented as median with interquartile range and Mann Whitney U test used for significance testing. Logistic regression was applied to explore risk factors associated with the outcomes and route of acquisition. To build logistic regression model forward selection was used and variables with p<0.10 were entered in the model. Results were not adjusted for BAME, because of unevenly distributed missing data. Likelihood ratio test was used to determine significant effect on the outcome after adjusting for the other variables in the model. Kaplan Meier plots were utilised to assess mortality.

Correlations between HA-COVID-19 and hospital bed occupancy, delayed RNA positivity, community incidence and staff self-reported COVID-19 sickness were explored by Pearson correlation coefficients.

All data was collated in excel and analysed in STATA using version 16.

## Results

On 25th Feb 2020 the first case of COVID-19 was recorded in the hospital. Subsequently until 18^th^ April, 2020, approximately 5000 people were tested, of which 1729 tested positive (see fig 1). Of these, 865 (50%) were admitted to hospital within 14d of testing for at least an overnight stay or already admitted to hospital at the time of testing. Due to the possibility of multiple acquisition sites, patients who had a separate hospital admission spell within 14d prior to testing (n=32) were excluded as were patients < 18 years of age (n=9) and those where no clinical data was available (n=3). Other categories are described in figure 1.

**Figure 1:**
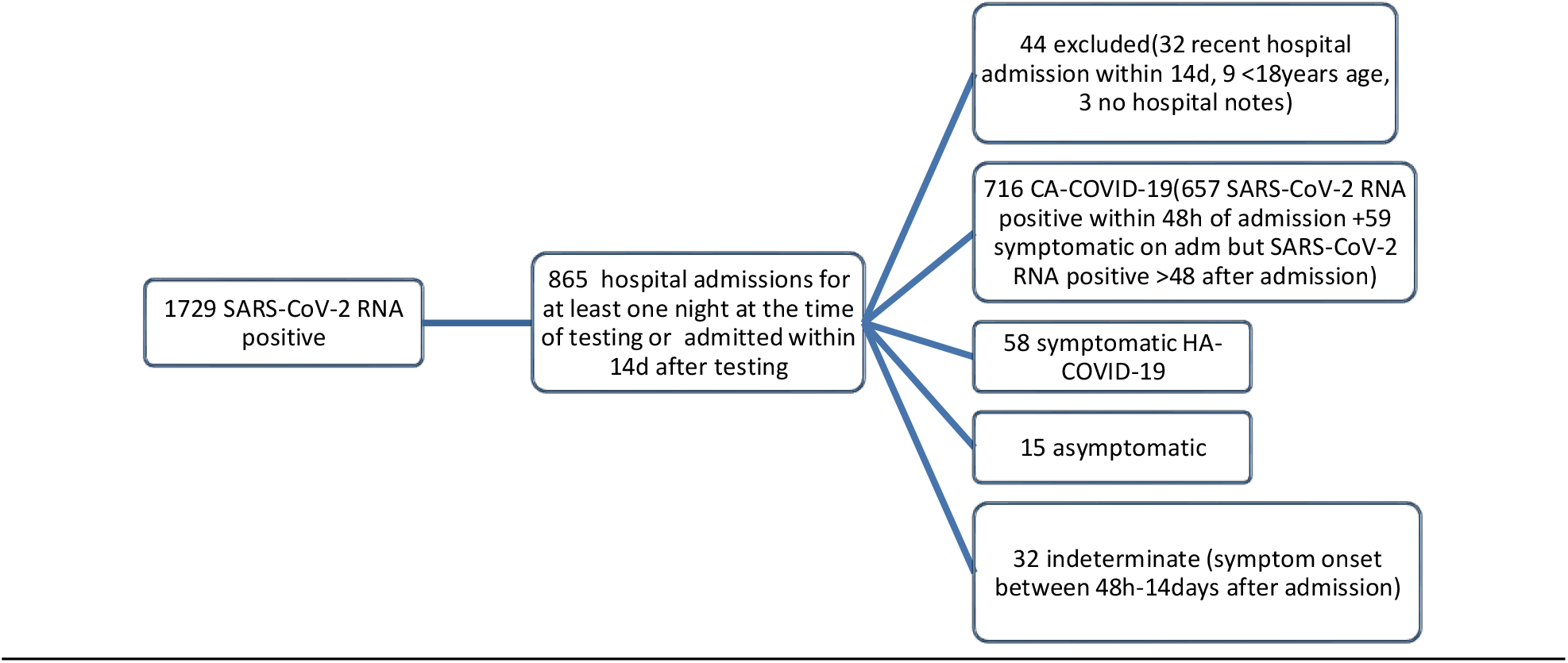
Classification of COVID-19 cases based on acquisition.

A total of 775 (82.8%) admitted cases were classified as CA-COVID-19, 58 (7.1%) as HA-COVID-19 and 32(3.7%) as indeterminate. Fifteen (1.7%) patients was classed separately as asymptomatic (2 SARS-CoV-2 RNA positive 48h to 7 days of admission, 2 cases 48h-7 days post admission and 11 cases post 14d of admission). For HA-COVID-19, time from admission to symptom onset ranged from 15 to 250 days (median 32.5 and IQR 21-65 days). Figure 2 illustrates the primary reason for admission of HA-COVID-19 and indeterminate cases. Incidence of HA-COVID-19 during this period was 133 per 100,000 bed-days.

**Figure 2:**
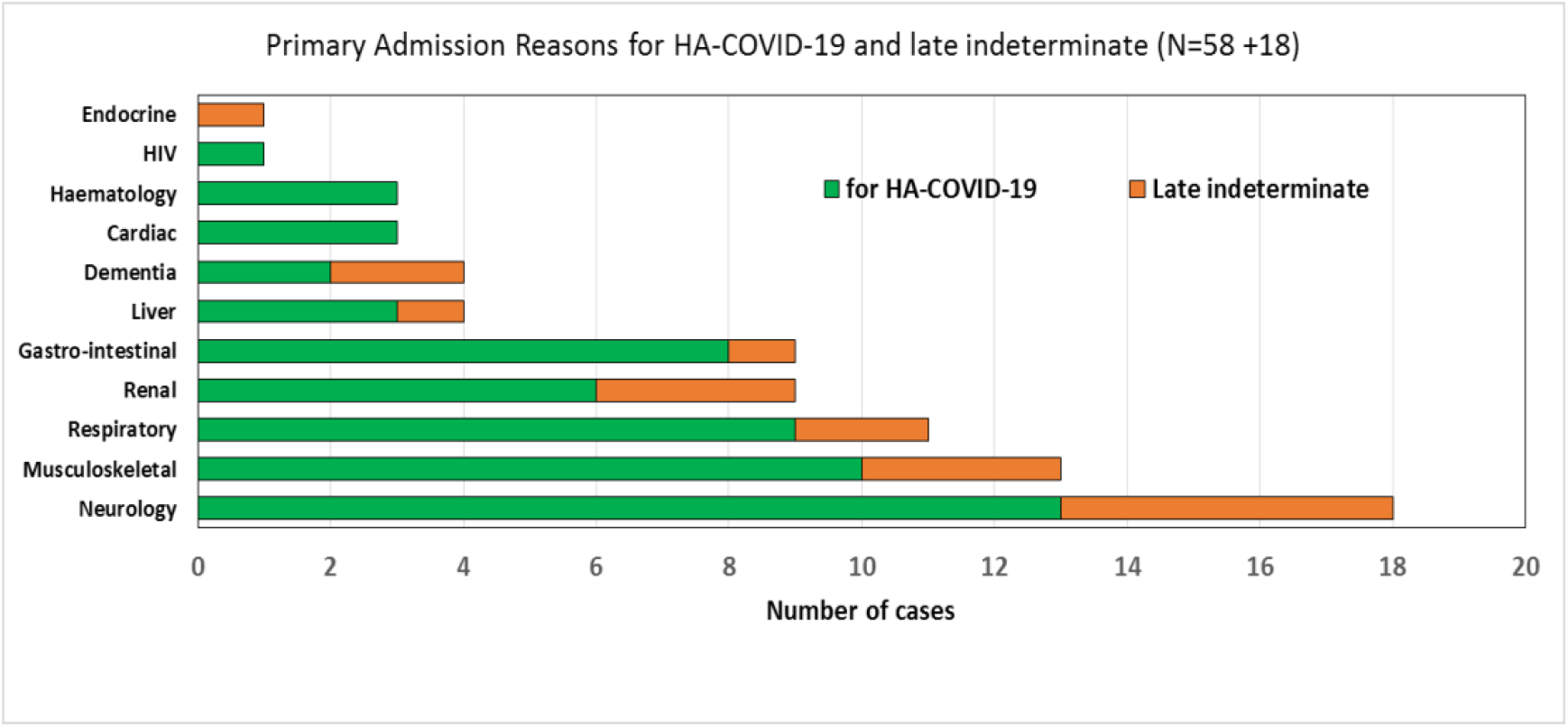
Primary admission diagnosis of HA-COVID-19 and late indeterminate cases.

Table 1A and 1B summarise the clinical features and outcomes of CA- and HA-COVID-19. Overall, the HA-COVID-19 population was more likely to be >65years and have a CCI ≥5, but less likely belong to BAME. Diabetes, CKD and malignancy were the individual comorbidities more common with HA-COVID-19 and a significantly longer post-COVID-19 length of stay (median 28d, p<0.001) was observed but no overall difference in mortality (also figure S1).

**Table 1A:** Risk factors and outcomes of patients with HA-COVID-19 vs CA-COVID-19.

| Overall Risk factors | CA-COVID-19 N=716 | % | HA-COVID-19 N=58 | % | Total N=774 | p-value |
| --- | --- | --- | --- | --- | --- | --- |
| Male | 422 | 58.9 | 29 | 50 | 451 (58.3%) | 0.184 |
| > 65 years | 353 | 49.3 | 38 | 65.5 | 391 (50.2%) | 0.018 |
| BAME* | 434 | 67.2 | 19 | 33.9 | 453 (64.5%) | <0.001 |
| Not BAME | 212 | 32.8 | 37 | 66.1 | 249 (35.5%) |  |
| Dementia | 83 | 11.6 | 10 | 17.2 | 93 (12.2%) | 0.203 |
| Hypertension | 407 | 56.8 | 32 | 55.2 | 439 (56.3%) | 0.805 |
| COPD/Asthma | 229 | 32.0 | 19 | 32.8 | 248 (32.0%) | 0.903 |
| Malignancy | 61 | 8.5 | 19 | 32.8 | 80 (10.3%) | <0.001 |
| CKD | 142 | 19.8 | 19 | 32.8 | 161 (20.8%) | 0.02 |
| Diabetes | 159 | 22.2 | 20 | 34.5 | 179 (23.1%) | 0.033 |
| CCI≥5 | 292 | 40.8 | 37 | 63.8 | 329 (42.5%) | <0.001 |
| Overall outcomes |  |  |  |  |  |  |
| 7 day mortality | 104 | 14.5 | 4 | 6.9 | 108 (14.0%) | 0.107 |
| 14 day mortality | 156 | 21.8 | 12 | 20.7 | 168 (21.6%) | 0.845 |
| 30 day mortality | 187 | 26.1 | 15 | 25.9 | 202 (26.1%) | 0.966 |
| Discharged within 30d | 440 | 61.5 | 23 | 39.7 | 463 (59.8%) | 0.001 |
| Median LOS survivors after COVID-19 diagnosis | 9 (IQR: 5-20) |  | 28 (IQR: 14-30) |  |  | <0.001 |
| ICU admission within 30d of diagnosis | 232 | 32.4 | 13 | 20.41 | 245(31.7%) | 0.116 |
| ICU admission: first 7 days of detection/symptoms onset | 221 | 30.9 | 12 | 20.7 | 233 (30.1%) | 0.104 |
| How many of the discharged patients have been readmitted within 30d n=464 | 42 | 9.6 | 2 | 8.7 | 44 (9.5%) | 0.892 |

**\*BAME: unknown in 72 patients**

**Table 1B:** Univariate and multivariate analysis for outcomes.

| Variables | Univariate |  |  | Model 1 |  |  | Model 2 |  |  |
| --- | --- | --- | --- | --- | --- | --- | --- | --- | --- |
|  |  |  |  | Multivariate |  |  | Multivariate |  |  |
| Outcome: Mortality | OR | 95%CI | p-value | OR | 95%CI | p-value | OR | 95%CI | p-value |
| HA-COVID-19 | 0.99 | 0.53-1.81 | 0.966 | 0.78 | 0.41-1.47 | 0.438 | 0.80 | 0.42-1.52 | 0.498 |
| CCI≥5 | 3.59 | 2.56-5.0 | <0.001 | 3.69 | 2.62-5.2 | <0.001 |  |  |  |
| Male | 1.84 | 1.31-2.59 | <0.001 | 1.87 | 1.31-2.67 | <0.001 | 2.03 | 1.42-2.91 | <0.001 |
| > 65 years | 3.48 | 2.45-4.9 | <0.001 |  |  |  | 3.29 | 2.28-4.73 | <0.001 |
| CKD | 2.35 | 1.63-3.40 | <0.001 |  |  |  | 1.80 | 1.22-2.67 | 0.003 |
| Outcome: Discharged within 30 days | OR | 95%CI | p-value | OR | 95%CI | p-value | OR | 95%CI | p-value |
| HA-COVID-19 | 0.41 | 0.24-0.72 | 0.011 | 0.44 | 0.25-0.78 | 0.005 | 0.44 | 0.25-0.78 | 0.005 |
| CCI≥5 | 0.47 | 0.35-0.63 | <0.001 | 0.50 | 0.37-0.67 | <0.001 |  |  |  |
| Male | 0.59 | 0.43-0.79 | <0.001 | 0.57 | 0.41-0.77 | <0.001 | 0.56 | 0.41-0.76 | <0.001 |
| > 65 years | 0.53 | 0.40-0.71 | <0.001 |  |  |  | 0.60 | 0.43-0.81 | <0.001 |
| CKD | 0.54 | 0.38-0.77 | <0.001 |  |  |  | 0.68 | 0.47-1.00 | 0.048 |
| Diabetes | 0.55 | 0.39-0.78 | <0.001 |  |  |  | 0.68 | 0.48-0.98 | 0.036 |
| Outcome: ICU admission | OR | 95%CI | p-value | OR | 95%CI | p-value | OR | 95%CI | p-value |
| HA-COVID-19 | 0.6 | 0.32-1.14 | 0.116 | 0.76 | 0.39-1.46 | 0.409 | 0.78 | 0.40-1.51 | 0.461 |
| CCI≥5 | 0.4 | 0.31-0.61 | <0.001 | 0.43 | 0.31-0.60 | <0.001 |  |  |  |
| Male | 2.1 | 1.48-2.84 | <0.001 | 2.13 | 1.53-3.00 | <0.001 | 2.05 | 1.47-2.85 | <0.001 |
| > 65 years | 0.4 | 0.26-0.50 | <0.001 |  |  |  | 0.40 | 0.28-0.55 | <0.001 |
| CKD | 0.4 | 0.29-0.68 | <0.001 |  |  |  | 0.59 | 0.38-0.92 | 0.021 |

Among the 245 patients admitted to ICU (see table S1A and S1B), risk factors were similar as above and there were no statistically significant differences in any of the calculated outcome measures.

Table S2 summarises the performance a single CTNS taken within 48h to detect CA-COVID-19 for use in patient placement. Overall, sensitivity was 92.2% (95% CI 92.9-93.6) and specificity 100% but negative predictive value 60.3% (57.3-63.3%).

A delayed RNA positive result was seen in 53 patients (see table 3A). In 14 cases the cause was delay in sampling and in 39 cases samples was taken within 48h but SARS-CoV-2 RNA was negative. In these patients age >65, non BAME ethnicity, diabetes and malignancy, CCI>5 were more common as was 30d mortality (p=0.01). This association remained significant in the multivariable model (see table 3B). 45 cases (85%) were not isolated appropriately as a result of the negative RNA test.

**Table 3A:** **Risk factors and outcomes of patients who had delayed SARS-CoV-2 RNA positivity.**

|  | Delayed RNA detection (n=53) | % | No delay in RNA detection (n=753) | % | Total (N=806 ) | p-value |
| --- | --- | --- | --- | --- | --- | --- |
| Age>65 years | 36 | 67.9 | 381 | 50.60 | 417 (52.7 %) | 0.015 |
| Male | 30 | 56.6 | 433 | 57.50 | 463(57.4%) | 0.898 |
| BAME* | 16 | 33.33 | 450 | 65.98 | 466 (63.8%) | 0.001 |
| Dementia | 9 | 16.98 | 89 | 11.82 | 98 (12.2%) | 0.266 |
| Hypertension | 35 | 66.00 | 427 | 56.71 | 462(57.3%) | 0.184 |
| Diabetes | 26 | 49.06 | 169 | 22.40 | 195 (24.2%) | <0.001 |
| Asthma/COPD | 18 | 33.96 | 241 | 32.01 | 259 (32.1%) | 0.768 |
| Malignancy | 12 | 22.64 | 76 | 10.09 | 88 (10.9%) | 0.005 |
| CKD | 16 | 30.20 | 156 | 20.70 | 172 (21.3%) | 0.104 |
| CCI≥5 | 30 | 56.60 | 326 | 43.29 | 356 (44.1%) | 0.059 |
| Outcome |  |  |  |  |  |  |
| 7 day mortality | 1 | 1.89 | 108 | 14.34 | 109 (13.5 %) | 0.01 |
| 14 day mortality | 8 | 15.09 | 166 | 22.05 | 174 (21.6%) | 0.235 |
| 30 day mortality | 22 | 41.51 | 191 | 25.37 | 213 (26.4%) | 0.01 |
| Discharged | 20 | 37.74 | 459 | 60.96 | 479 (59.4%) | 0.001 |
| How many of the discharged patients have been readmitted within 30d- n=479 | 2 | 10.00 | 44 | 9.59 | 46 (9.6%) | 0.951 |
| ITU admission | 18 | 33.96 | 236 | 31.34 | 254 (32.5%) | 0.691 |
| ITU admission < 7 days | 16 | 30.20 | 224 | 29.60 | 240 (29.8%) | 0.946 |
| Median length of Stay | 24 (IQR: 15-30) |  | 10 (IQR: 5-22) |  | 11 (IQR: 5-23) | <0.001 |

\*BAME unknown in 76 patients

**Table 3B:** **Univariate and multivariate analysis for factors associated with Delayed SARS-CoV-2 RNA positivity and mortality**

| Variables | Univariate |  |  | Model 1 |  |  | Model 2 |  |  |
| --- | --- | --- | --- | --- | --- | --- | --- | --- | --- |
|  |  |  |  | Multivariate |  |  | Multivariate |  |  |
| Outcome:<br>Mortality | OR | 95%CI | p-value | OR | 95%CI | p-value | OR | 95%CI | p-value |
| Delayed SARS-CoV-2 RNA positivity | 2.088 | 1.18-3.70 | 0.01 | 1.91 | 1.05-3.50 | 0.034 | 1.78 | 0.99-3.25 | 0.056 |
| CCI≥5 | 3.542 | 2.54-4.93 | <0.001 | 3.58 | 2.55-5.0 | <0.001 |  |  |  |
| Male | 1.897 | 1.36-2.64 | <0.001 | 2.01 | 1.42-2.83 | <0.001 | 2.17 | 1.53-3.07 | <0.001 |
| > 65 years | 3.178 | 1.26-2.46 | <0.001 |  |  |  | 2.64 | 1.83-3.81 | <0.001 |
| CKD | 2.28 | 1.59-3.26 | <0.001 |  |  |  | 1.6 | 1.09-2.36 | 0.017 |
| Hypertension | 1.619 | 1.13-2.29 | 0.007 |  |  |  | 1.56 | 1.08-2.26 | 0.019 |

A potential source patient was found for 44 HA-COVID-19, 14 late indeterminate and 11 asymptomatic late indeterminate (see table 4). CA-COVID-19 with a delayed RNA detection contributed to the majority of HA-COVID-19 and late indeterminate cases (34.5 %).

**Table 4:** Potential patient sources for HA-COVID-19 and late indeterminate cases.

| Source patient | No. of HA-COVID-19<br>N=44* | No. of late indeterminate cases<br>N=14** | Asymptomatic (late indeterminate)<br>N=11*** | % (HA-COVID-19 + late indeterminate)<br>N=58 |
| --- | --- | --- | --- | --- |
| CA-COVID-19 | 4 | 3 | 1 | 12.07 |
| CA-COVID-19 delayed diagnosis | 14 | 6 | 0 | 34.48 |
| HA-COVID-19 | 15 | 3 | 7 | 31.03 |
| Indeterminate (total) | 11 | 2 | 3 |  |
| Late indeterminate | 8 | 2 | 2 | 17.24 |
| Early indeterminate | 3 | 0 | 1 | 5.17 |

\* Potential patient source not established for 14 patients
\*\* Potential patient source not established for 4 patients
\*\*\*Potential patient source not established for 2 patients

Figure S2A represents the incidence of self-reported staff sickness from 2019 and 2020 and figure S2B represents the self-reported COVID-19 sickness absence for groups with high (G1 and G2) and low (G3) risk of nosocomial exposure.

Bed occupancy varied during the study period as seen in figure S3, with an early fall in bed occupancy reflecting the reduction of elective activity and expedited discharge of non-COVID-19 patients in anticipation of increasing COVID-19 admissions.

Correlation between weekly incidence of HA-COVID-19 (including late indeterminate cases) and staff self-reported sickness absence, delayed RNA positive cases, community incidence and Trust COVID-19 bed occupancy is displayed in figure 3. Significant correlation was observed with the former two but neither with COVID-19 bed occupancy nor the incidence in the community.

**Figure 3:**
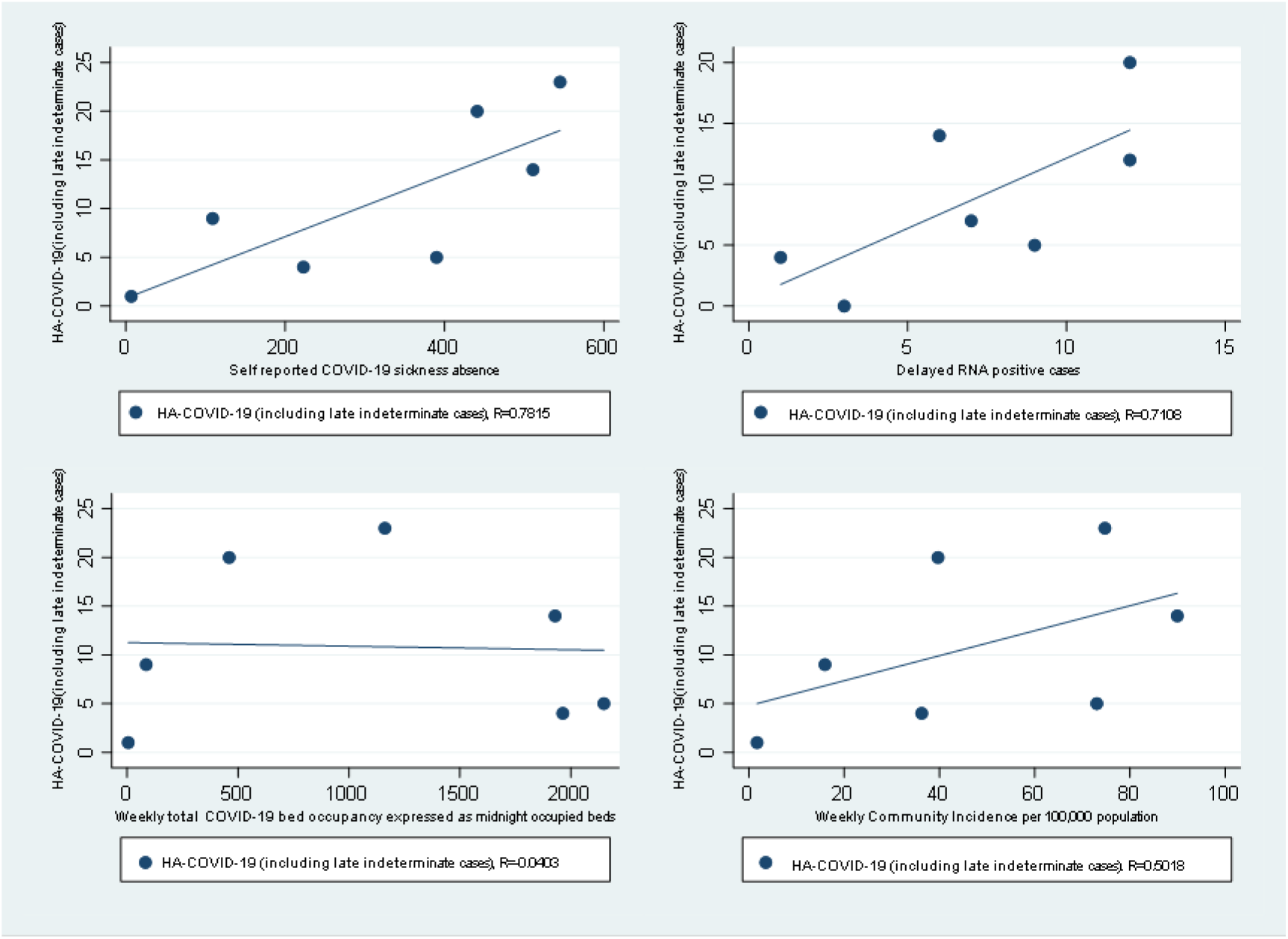
Correlation between weekly incidence of HA-COVID-19 (including late indeterminate cases) and staff self-reported sickness absence, delayed RNA positive cases, community incidence and COVID-19 bed occupancy

The incidence of HAB during the study period was comparable to the average of the previous two years (see figure S4).

A 5.12% difference was observed when symptomatic classification was compared with SARS-CoV-2 RNA detection based classification (including 11 cases of symptomatic CA-COVID-19 classified as HOPHA and 1 as HODHA (see supplement table S6)).

Tables S2-S4 (supplementary data) provide risk factors and outcome analysis including the indeterminate cases. Table S5 provides additional outcome measures for Delayed SARS-CoV-2 RNA positivity. Table S7 and figures S6 provide the estimated incubation period for HA-COVID-19 (N=44 patients).

## Discussion

A recent survey from 46 acute hospitals in the UK reported an average of approximately 8-9% of patients with a positive COVID-19 whose diagnosis was identified 14d after admission (inter-quartile range 3.8% to 12%) (written communication). In the present report 7.1% were symptomatic HA-COVID-19 and an additional 2.2% were symptomatic late indeterminate cases. In addition, 11 asymptomatic cases were identified after 14d and 2 were identified 8-14d from admission. We identified asymptomatic cases as part of contact screening in high transmission risk situations but if outbreak management programmes include consistent testing of all asymptomatic contacts, potentially more cases can be identified.

Both in the overall analysis and in those admitted to ICU distinct differences in the risk factors between CA-COVID-19 and HA-COVID-19 were observed. The older and more comorbid HA-COVID population likely represent the overall hospitalised population at risk of HAI. However the lower 7d mortality (14.5% vs 6.9%) and lower proportion admitted to ICU (32.4 vs 20.4%) may be because CA-COVID-19 cases were admitted to hospital only if cases were severe enough to warrant admission. Overall 30d mortality appears similar between the two groups. We also observed a trend towards HA-COVID-19 cases of BAME origin (67.2% vs 33.9% p<0.001) but since data was missing for 72 patients, results are not conclusive.

In our setting, the sensitivity of single CTNS taken within 48h to predict the potential to transmit SARS-CoV-2 was 92.2% and the negative predictive value was 60.3%. Wang et al (11) highlighted the variability in detection of the viral RNA by PCR in different sample types with maximum positivity rate in bronchoalveolar lavage (93%), followed by sputum (72%) and nasal swabs (63%).

In the present study, 53 patients had a delayed virological diagnosis in keeping with the recent report of Kucirka et.al (12) who reviewed the variation in false negative SARS-CoV-2 RT PCR results from upper respiratory tract samples. They concluded that the probability of a false-negative result in an infected person varied with days from symptom onset. On the day of symptom onset, the median false-negative rate was 38% (CI, 18% to 65%) which decreased to 20% (CI, 12% to 30%) on day 8 (3 days after symptom onset) then began to increase again, from 21% (CI, 13% to 31%) on day 9 to 66% (CI, 54% to 77%) on day 21. Since patients can present at any stage of the illness, one CTNS would seem insufficient to prevent onward transmission if the decision to segregate patients is based on this result alone. Gao et al (5) have described two nosocomial outbreaks in Wuhan where the index was a misdiagnosed case of CA-COVID-19. Our results also suggest that 34.5% of all HA-COVID-19 and late indeterminate infections could be traced back to cases where the acquisition was in the community but an RNA based diagnosis could not be made within 48h of admission (table 5). The weekly incidence of delayed RNA positivity also correlated with the incidence of HA-COVID-19.

Among those patients where a virological diagnosis was delayed, co-morbidities (CCI>5) were higher than in those without a delay (56.6% vs 43.3% p=0.058) and may explain the difficulty in making a clear diagnosis in the presence of multiple clinical features. However, the higher 30d mortality (p=0.01) (table 4B) in this patient group emphasises the need identify a more accurate method of ruling out COVID-19 in the initial stages of presentation. A combination of detailed history taking, successive swabs, deeper respiratory sample and radiology and biochemical markers should be evaluated in the future to help determine the best strategy to reduce onward transmission in hospital settings.

During Feb-April 2020, staff sickness due to cold, cough, flu and chest and respiratory problems decreased from the second week of the study probably due to the similarity of symptoms with COVID-19 and the introduction of a new staff sickness code (S13 COVID-19 see figure S2A). COVID-19 sickness on the other hand, increased in the first three weeks of the study and then started decreasing from week 4 which coincides with the introduction of community wide restriction to movement and closure of non-essential businesses. This finding is similar to with the recent report by Hunter et al (13) which found an increase in staff positivity from 5 to 20% from 10^th^ to 31^st^ March 2020. They compared positivity rates amongst staff in patient-facing, non-patient facing and non-clinical roles and found no significant difference between these groups suggesting nosocomial transmission from patients to staff was not an important factor during the study period. However in their study, data on clinical roles was only available for 1/3 of included staff. Our study compared staff self-reported COVID-19 sickness rates between patient facing high nosocomial risk and non-clinical staff and found the difference increased after social distancing was implemented. We hypothesize that working for home may be an easier option for non-patient facing staff that may not need to take sickness leave even if mild symptoms are present.

Previous work has shown that healthcare staffing potentially influences the incidence of healthcare-associated infections (14). In the present study, the incidence of HA-COVID-19 correlated positively with healthcare staff absence due to COVID-19. The reasons for this correlation are likely to be multifactorial since the reduction in healthcare staff to patient ratios may have a negative influence on appropriate IPC measures but may independently be a reason for SARS-CoV-2 transmission from infected healthcare staff to patients.

Dona et al (15) recently discussed the potential impact of COVID-19 on hospital transmission of MDR organisms, based on how risk factors such a healthcare absence, hospital overcrowding, PPE usage and patient demographics are distributed in a healthcare setting. In our study, the effect of the measures taken and the demographics did not have a significant impact on the rate of MDR healthcare-associated infections when compared to the average of the previous two years.

Limitations of our study: Staff members were tested for SARS-CoV-2 via a regulated pathway from 27th March 2020. Prior to this, testing was on a special request only and has not been included in this report. Due to the high community prevalence at the time it was not possible to determine the source of the infection linked to staff members. Recent reports suggest SARS-CoV-2 can be transmitted by asymptomatic carriers (16, 17). We have observed this in HA-COVID-19 clusters but this study did not extend to include detection of asymptomatic infections. It is possible our HA-COVID-19 rates are lower during this period because a large proportion of elective work had stopped which limited the number of susceptible patients in the hospital.

## Recommendations, challenges and future directions

This study shows that hospital transmission of COVID-19 can be initiated by carriers who may not show symptoms, and could be admitted for other reasons. Screening for asymptomatic or early infection on admission is one approach recently advocated to segregate COVID-19 and non COVID-19 patients. However the use of single CTNS for this purpose is limited. Further work on appropriate use of resources in patient pathways to limit transmission is recommended.

## Data Availability

not available

## Acknowledgements

Informatics Team in BIU: Christopher Fry, Dominic Thurgood and Isuf Ali for assistance with extracting electronic data from various Trust IT systems.

Workforce Development Team: Christopher Oram for assistance with extracting staff sickness records.

Occupational health team: Helen Parsons

KCH Infection Prevention and Control nurse’s team Viapath Infection Sciences

